# Validation of a Tissue-Based Predictive RNA Test for Immunotherapy Benefit in NSCLC: the PREDAPT study

**DOI:** 10.64898/2025.12.12.25342169

**Authors:** Kevin C. Flanagan, Jon Earls, Jeffrey Hiken, Rachel L. Wellinghoff, Michelle Ponder, Kyle Pemberton, Orlan K. Macdonald, Karim Welaya, Andrew W. Pippas, Karl D’Silva, Xingwei Sui, Warren L. Alexander, Jennifer Slim, Steven J. Saccaro, Todd D. Shenkenberg, Samuel D. Bailey, Scott A. Sonnier, Georges Azzi, Bruce Bank, Steven E. Kossman, Paul Gonzales, James Wade, Jessica A. Hellyer, Howard L. McLeod, Eric J. Duncavage, Jarret I. Glasscock

## Abstract

Lung cancer remains the leading cause of cancer mortality worldwide. Immune checkpoint inhibitors targeting PD-1/PD-L1 have significantly improved outcomes in a subset of patients. OncoPrism, a clinical test employing a multidimensional predictive RNA-based immune biomarker, was evaluated for predicting immune checkpoint inhibitor (ICI) benefit in non-small cell lung cancer (NSCLC) patients. This study included data from 1,487 patients and evaluated OncoPrism across four NSCLC cohorts: one PD-L1 inhibitor cohort (n=195), one PD-1 inhibitor cohort (n=89), and two non-ICI cohorts (n=193 and n=1,010, respectively). In the PD-L1 inhibitor cohort, OncoPrism predicted progression-free survival (p<0.0001) and overall survival (p=0.043). In the PD-1 inhibitor cohort, an observational clinical trial, PREDAPT (NCT04510129) enrolling patients from 17 healthcare systems, OncoPrism predicted overall response rate (p=0.008), progression-free survival (p=0.004), and overall survival (p=0.011). PD-L1 Tumor Proportion Score (TPS) was not predictive of response, progression-free survival, or overall survival. OncoPrism did not predict overall survival across two non-ICI NSCLC cohorts (p=0.54, p=0.73), suggesting the test is specifically predictive of ICI benefit rather than being prognostic with more limited clinical utility. Overall, the data show OncoPrism high patients are likely to benefit from a two to three-fold increase in overall response rate, progression-free survival, and overall survival compared to those in other OncoPrism groups. These results underscore the impact of OncoPrism to address the current unmet need for ICI response prediction in NSCLC.

## INTRODUCTION

Non-small cell lung cancer (NSCLC) affects over 230,000 individuals annually in the United States, resulting in around 127,000 deaths.^1,2^ The nearly half of all patients diagnosed at stage IV have a dire prognosis.^3,4^ The five-year overall survival (OS) rate for NSCLC is around 27%, with the five-year relative survival rate for patients with distant metastases being just 9%.^2,5^

Current treatment options for late-stage NSCLC diagnosis include surgical intervention, radiation therapy, chemotherapy, and targeted drug therapies. The introduction of immune checkpoint inhibitors (ICI), such as anti-PD-1 and anti-PD-L1 antibodies, has transformed the therapeutic landscape, approximately doubling five-year OS versus platinum-based chemotherapy in some contexts.^6–8^ However, only a minority of ICI-treated patients achieve durable benefit, underscoring the urgent need for precision medicine approaches that identify the subset of NSCLC patients most likely to benefit from ICI.

Despite considerable effort over the past decade, progress toward effective ICI predictive biomarkers in NSCLC has been limited.^9,10^ PD-L1 Tumor Proportion Score (TPS) remains the only predictive clinical biomarker consistently recommended to guide ICI treatment decisions in NSCLC patients across leading treatment guidelines (NCCN, ASCO, ESMO).^11–13^ Unfortunately, PD-L1 TPS has only modest positive predictive value, often resulting in escalation of treatment to ICI plus chemotherapy regimens. Despite significant efforts in the field, other potential biomarkers have shown limited clinical utility. Mutations in genes such as *STK11* and *KEAP1* are associated with poor response to ICI, but their utility as predictive rather than prognostic markers is not clear.^14^ High specificity predictive biomarkers positively associated with ICI response are lacking. Microsatellite instability-high (MSI-H) is rare in NSCLC (<1%).^15^ Despite initial promise, tumor mutational burden (TMB) has been diminished from national guidelines due to modest correlation with response, issues of assay standardization, and inconsistent TMB-high cutoffs.^16^ As a result, clinicians are left with PD-L1 TPS as the sole tool to predict response to ICI.

To make precision medicine a reality for NSCLC patients facing ICI therapy decisions, new predictive biomarkers must improve accuracy with balanced improvement in both sensitivity and specificity beyond PD-L1 TPS. New biomarkers must also demonstrate robust analytical performance across commonly encountered pre-analytic variables in clinical testing. Furthermore, such tests must be compatible with limited formalin-fixed, paraffin-embedded (FFPE) tissue specimens to work with existing clinical workflows and enable widespread adoption. Finally, predictive biomarkers of ICI response must show clinical specificity rather than simply being prognostic.^17^ Predictive biomarkers are specific to the benefit from one class of therapy whereas prognostic biomarkers are predictive of outcome regardless of therapy. Unlike prognostic biomarkers, predictive biomarkers are clinically actionable to improve patient outcomes.^18–20^

Previously, we reported the clinical and analytical validation of OncoPrism-HNSCC, a test that addresses the ICI-predictive diagnostic needs detailed above in head and neck cancers. OncoPrism-HNSCC is a robust novel ICI predictive diagnostic employing RNA, machine learning, and an immune-based multi-dimensional biomarker capable of predicting clinical benefit from ICI in recurrent or metastatic head and neck squamous cell carcinoma (RM-HNSCC).^21,22^ This test uses RNA expression data from the immune microenvironment of FFPE tumor samples to categorize patients based on likelihood of disease control in response to ICI with high sensitivity and specificity. OncoPrism is a unique, predictive classifier that, at its core, is built from expression signatures of immune-related genes with observed expression across numerous cancers. Therefore, we hypothesized that OncoPrism could predict ICI response in indications beyond RM-HNSCC. We sought to evaluate whether the OncoPrism biomarker predicts response in NSCLC. Here, we report the tuning and validation of an NSCLC-specific test, OncoPrism-NSCLC, using the same underlying biomarker described previously.

## RESULTS

### Establishing an NSCLC ICI Prediction Model

The distribution of OncoPrism scores varied across cancer indications (Figure 1A). Therefore, a new distribution was created using 1,010 NSCLC patient samples to tune the biomarker underlying OncoPrism-HNSCC for use in NSCLC patients. This NSCLC-specific distribution set thresholds dividing the lower, middle, and high scoring tertiles of the patient population, resulting in OncoPrism low (0-40), medium (41-57), and high (58-100) groups (Figure 1B). These thresholds and resulting OncoPrism groups were used for all subsequent work to interrogate the correlation between OncoPrism group and clinical benefit from ICI.

**Figure 1.**
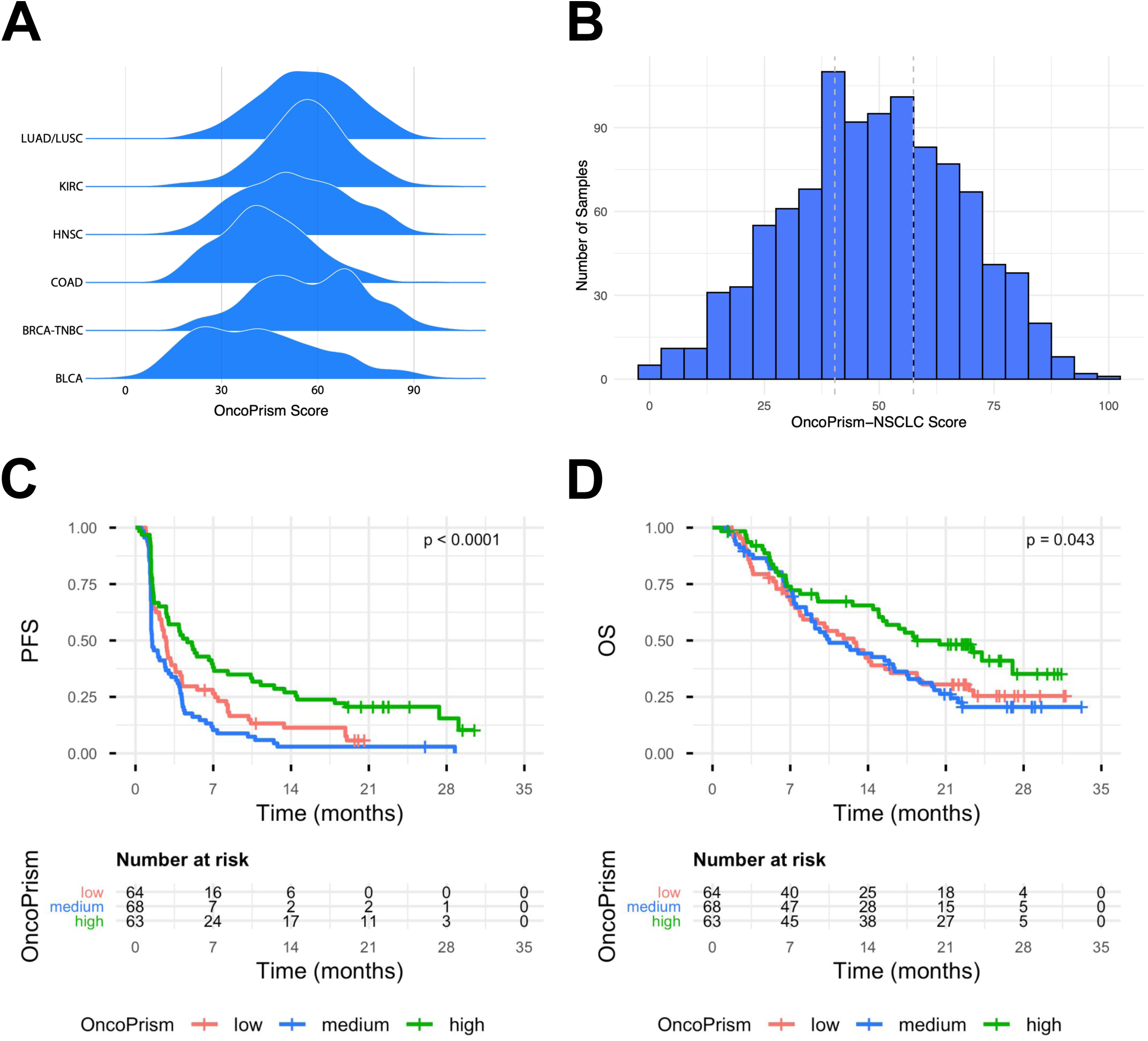
**(a)** The OncoPrism score distribution varies by indication. The distribution of OncoPrism scores, scaled relative to the entire dataset rather than the specific indication, is shown for TCGA data from six indications: lung adenocarcinoma/lung squamous cell carcinoma (LUAD/LUSC); kidney renal clear cell carcinoma (KIRC); head and neck squamous cell carcinoma (HNSC); colon adenocarcinoma (COAD); triple negative breast cancer, (BRCA-TNBC); and bladder carcinoma, (BLCA). **(b)** OncoPrism-NSCLC scores were scaled using TCGA LUAD and LUSC data. The 33rd and 67th percentile values were used as the thresholds between low (0-40), medium (41-57), and high (58-100) OncoPrism groups. **(c)** OncoPrism predicted progression-free survival (PFS) in patients treated with anti-PD-L1 therapy (OAK ICI cohort, n=195). **(d)** OncoPrism predicted overall survival (OS) in patients treated with anti-PD-L1 therapy (OAK ICI cohort, n=195).

### Validation of OncoPrism-NSCLC in PD-L1 Inhibitor Cohort

Having established an NSCLC-specific OncoPrism model, we evaluated its performance in a cohort of 195 Stage IIIB or IV NSCLC patients treated with anti-PD-L1 ICI after one or two previous chemotherapy regimens in the OAK trial.^23^ OncoPrism high patients had significantly longer progression-free survival (PFS; median=4.7 months) than OncoPrism low (median=1.5 months) and medium (median=2.8 months) patients (Figure 1C, p<0.001). Likewise, OS was longer for OncoPrism high patients (median=20.4 months) than low (median=12.7 months) and medium (median=10.5 months) patients (Figure 1D, p=0.043).

### Validation of OncoPrism-NSCLC in the Multicenter PREDAPT PD-1 Inhibitor Cohort

Given the predictive value of OncoPrism in the PD-L1 Inhibitor OAK cohort, we proceeded to evaluate the performance of OncoPrism-NSCLC in a multicenter, retrospective clinical trial, PREDAPT (NCT04510129). A total of 176 patients were enrolled from 17 sites at the time of this readout. Samples from 89 patients across 12 sites met all eligibility and quality control criteria and were included in the validation study (Figure 2; Table S1). Patient clinical characteristics were broadly similar to the 154 patients in the ICI arm of the KEYNOTE-024 trial comparing outcomes in pembrolizumab vs. chemotherapy treated patients (Table 1).^24^

**Figure 2.**
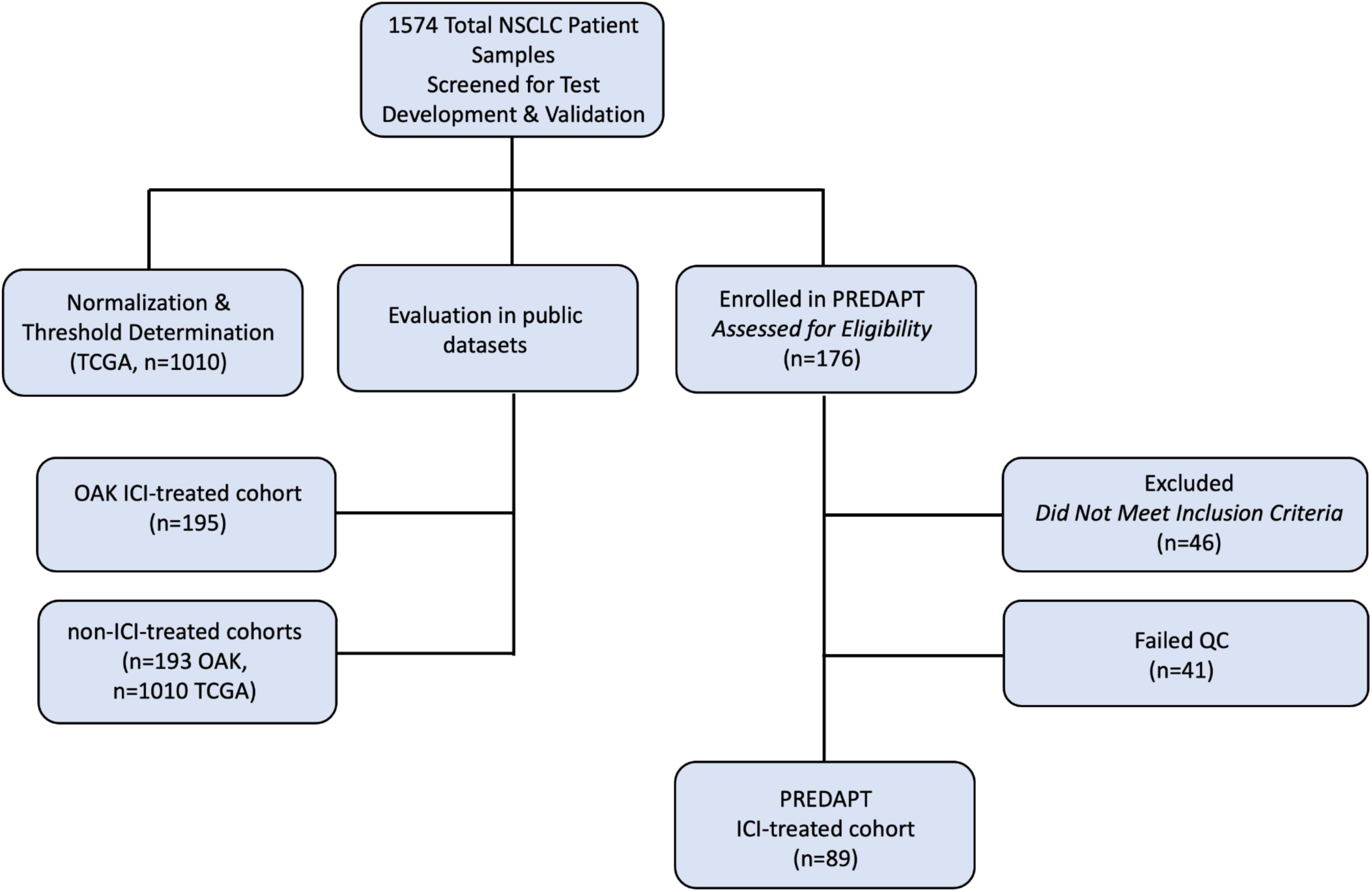
Study Overview. 1,574 patient samples were screened or used in this study. TCGA patient samples (n=1,010) were used to normalize the data and set NSCLC thresholds. The test was validated using 1,487 patient samples across four NSCLC cohorts: PD-L1 inhibitor-treated patients (OAK ICI cohort, n=195), PD-1 inhibitor-treated patients (PREDAPT ICI cohort, n=89), and two non-ICI cohorts (OAK and TCGA, n=193 and n=1,010, respectively). Of 176 samples assessed for inclusion in the PREDAPT trial, 46 were excluded for eligibility reasons and 41 failed quality control (QC).

**Table 1.** Clinical Data.

| Characteristic | Levels | PREDAPT | KEYNOTE-024 |
| --- | --- | --- | --- |
| Age (median) | Age | 67 | 64.5 |
| Sex | Male | 51 (57.3%) | 92 (59.7%) |
|  | Female | 38 (42.7%) | 62 (40.3%) |
| ECOG | Not available | 10 (11.2%) | - |
|  | 0 | 19 (21.3%) | 54 (35.1%) |
|  | 1 | 55 (61.8%) | 99 (64.3%) |
|  | 2 | 5 (5.6%) | 2 (0.6%) |
| Smoking Status | Used to smoke cigarettes | 50 (56.2%) | 115 (74.7%) |
|  | Currently smoke cigarettes | 22 (24.7%) | 34 (22.1%) |
|  | Not known | 11 (12.4%) | - |
|  | Never smoked cigarettes | 6 (6.7%) | 5 (3.2%) |
| Histology | Nonsquamous | 63 (70.8%) | 125 (81.2%) |
|  | Squamous | 26 (29.2%) | 29 (18.8) |
| Therapy | ICI plus chemo | 63 (70.8%) | - |
|  | Mono ICI | 26 (29.2%) | 154 (100%) |
| PD-L1 TPS | <1 | 37 (44.6%) | - |
|  | 1-49 | 18 (21.7%) | - |
|  | 50+ | 28 (33.7%) | 154 (100%) |

Because thresholds between OncoPrism groups were set using the tertile scores in the reference dataset, the expected distribution of patients was one-third of patients in each OncoPrism group. In the validation cohort, 28% of patients were in the low group, 40% of patients were in the medium group, and 31% of patients were in the high group. Median length of follow up was 9.3 months.

A significant trend towards higher overall response rate (ORR) was observed from the low to medium to high OncoPrism groups (p=0.0083; Figure 3A). The OncoPrism high ORR was 71%, compared to just 36% for both OncoPrism low and medium. The overall study ORR was 47%. Similarly, patients in the OncoPrism high group also had significantly longer PFS (median=18.6 months) than patients in the low and medium groups (median=7.0 & 5.5 months, respectively; p=0.0038, Figure 3C). Likewise, the higher ORR and PFS corresponded with significantly longer OS in OncoPrism high patients (median=22.9 months) relative to low (median=13.0) and medium (median=9.2) patients (p=0.011, Figure 3E).

**Figure 3.**
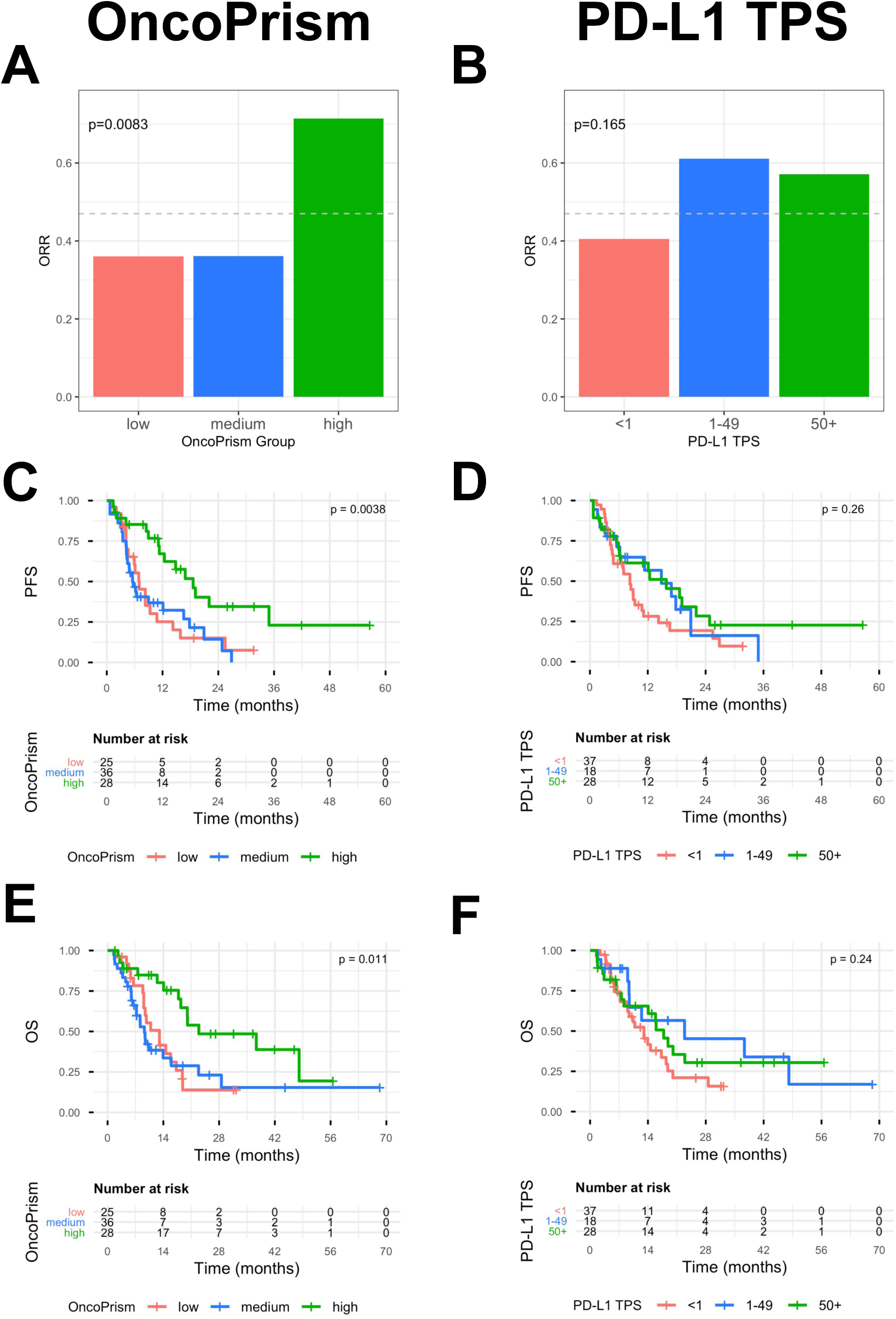
OncoPrism group, but not PD-L1 TPS, is correlated with clinical benefit. OncoPrism-NSCLC (n=89, left) and PD-L1 TPS (n=83, right) were evaluated for their ability to predict clinical benefit. **(a-b)** OncoPrism-NSCLC group was significantly correlated with overall response rate (ORR), while PD-L1 was not. The ORR for the entire study population is shown as a dashed line. **(c-d)** OncoPrism-NSCLC, but not PD-L1 TPS, significantly predicted progression-free survival (PFS). **(e-f)** OncoPrism-NSCLC, but not PD-L1 TPS, significantly predicted overall survival (OS).

### OncoPrism-NSCLC vs. PD-L1 TPS Predictive Measures

Next we compared the performance of OncoPrism-NSCLC to the performance of PD-L1 in the PREDAPT study. PD-L1 TPS results were not available for 6 patients, so they were excluded from the PD-L1 analysis. OncoPrism-NSCLC results for the remaining 83 patients are shown in Figure S1. The commonly used TPS categories of TPS<1, TPS 1-49, and TPS≥50 were used to group patients. Unlike OncoPrism-NSCLC (Figure 3A,C,E & Figure S1), PD-L1 TPS was not predictive of ORR, PFS, or OS (Figure 3B,D,F) in these 83 patients. OncoPrism-NSCLC also had numerically higher accuracy, sensitivity, specificity, positive predictive power and negative predictive value than PD-L1 TPS (Table S2).

To examine the cohort results at the individual patient level, we plotted all 89 patients, sorted by their OncoPrism group, response label, PD-L1 status, treatment regimen, and disease subtype (Figure 4A). We tested for significant associations among these variables using the chi-squared test of independence. There was a significant association between OncoPrism group and response (p=0.008), with an enrichment of responders in the OncoPrism high group. While the majority of these OncoPrism high true responders are also TPS≥50, both TPS<1 and TPS 1-49 patients were also represented among these OncoPrism high true responders (Figure 4). Overall, there was a significant but modest association between OncoPrism-NSCLC and PD-L1 TPS (p=0.006). PD-L1 TPS category was also associated with treatment type with more monotherapy patients among TPS≥50 patients (p<0.001). Finally, OncoPrism group was associated with disease histology with fewer squamous cancers among OncoPrism high patients (p=0.028).

**Figure 4.**
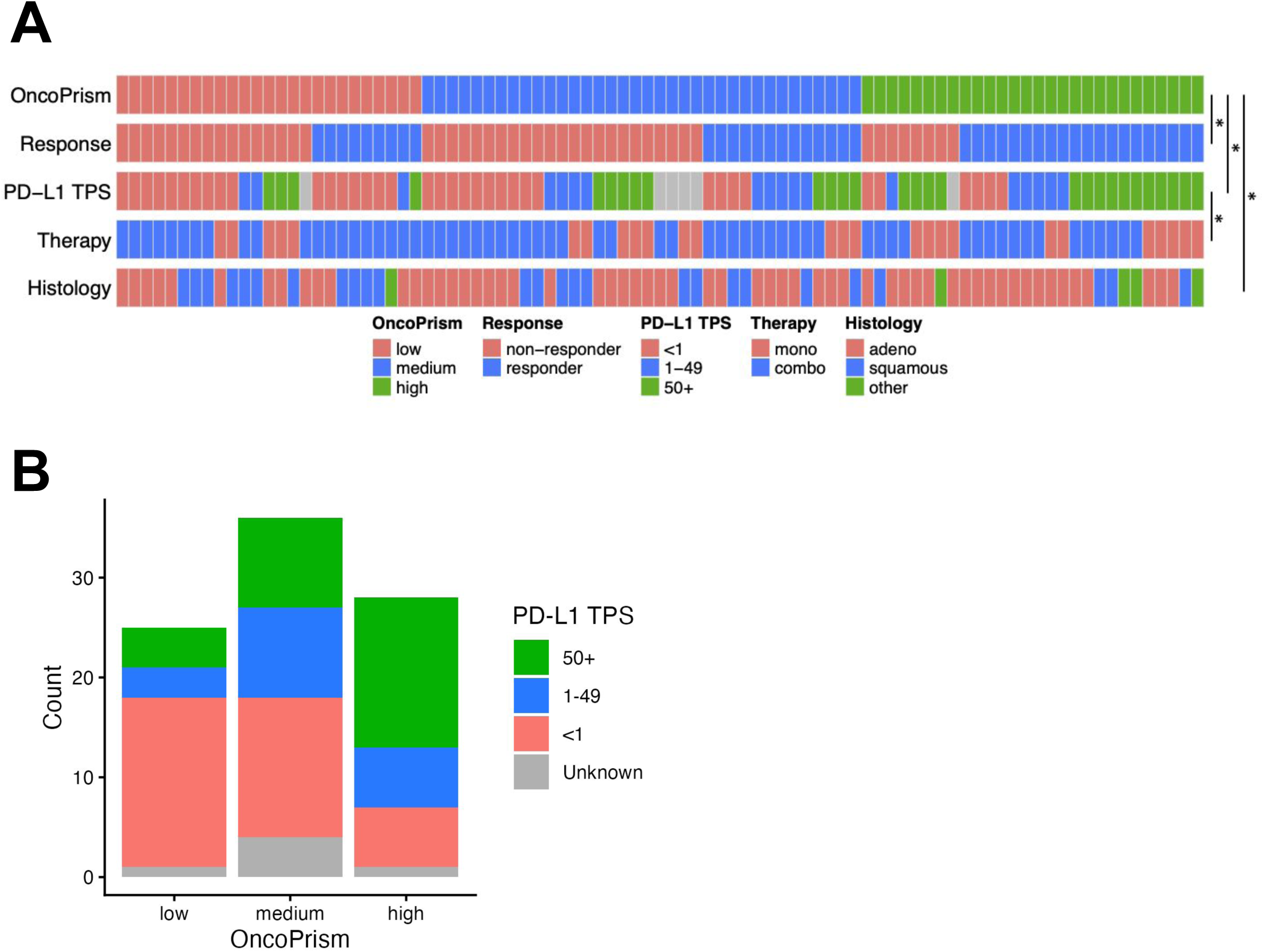
**(a)** Patient-level data for OncoPrism group, response, PD-L1 TPS category, therapy regimen, and histology type. Categorical variables with significant associations are noted (*p<0.05, chi-squared test of independence). **(b)** Number of PREDAPT patient samples in each PD-L1 TPS category (TPS<1, 1-49, ≥50, and unknown) by OncoPrism group.

Table S3 compares predictions and outcomes for OncoPrism-NSCLC and PD-L1 TPS. While OncoPrism-NSCLC and PD-L1 TPS were concordant for 58/83 results, for discordant results PD-L1 had both more false negatives (9 vs. 5) and more false positives (8 vs. 3) than OncoPrism-NSCLC. Responders were treated as the positive class and non-responders were treated as the negative class. Of the 22 responders in the OncoPrism-NSCLC low and medium groups (i.e., false negatives), only 3 were treated with monotherapy ICI. Likewise, only 2 of 26 PD-L1 TPS false negatives were treated with monotherapy ICI, suggesting that response to chemotherapy may be driving many of the false negatives for both OncoPrism-NSCLC and PD-L1 TPS.

### Assessing Predictive vs. Prognostic Characteristics of OncoPrism-NSCLC

To determine if OncoPrism-NSCLC was predictive of response to ICI or prognostic of outcome generally, we evaluated the performance of OncoPrism-NSCLC in patients who were not treated with ICI. OncoPrism-NSCLC did not predict PFS or OS in patients treated with docetaxel (OAK non-ICI, n=193, Figure 5A-B). Likewise, OncoPrism-NSCLC was not predictive of OS in the TCGA data, composed of patients treated with a variety of non-ICI therapies (TCGA non-ICI, n=1,010, Figure 5C).^25^ Together, these data indicate that OncoPrism-NSCLC is not broadly prognostic and may be predictive of ICI specifically.

**Figure 5.**
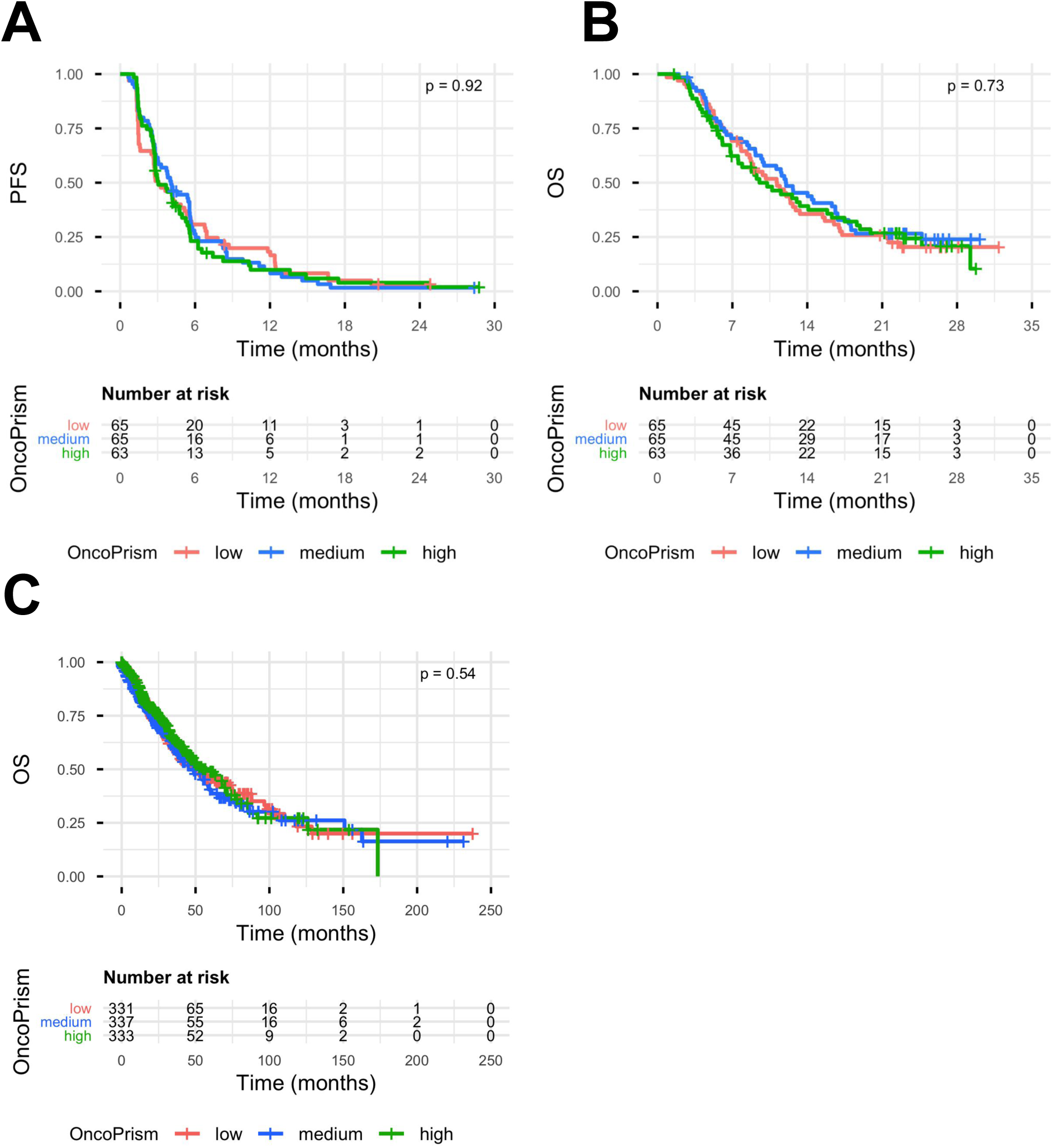
OncoPrism-NSCLC does not predict survival in non-ICI-treated patients. OncoPrism group was not correlated with **(a)** progression-free survival (PFS) or **(b)** overall survival (OS) in docetaxel-treated NSCLC patients (n=193). **(c)** OncoPrism-NSCLC was not predictive of OS in non-ICI treated patients from TCGA NSCLC data (n=1,010).

### Characterizing Robustness Through Assessment of Analytical Variation

To evaluate the analytical performance of OncoPrism-NSCLC, two operators processed three replicates each of seven different biological samples with a range of OncoPrism scores (23-75) on two different days using two different reagent lots (Figure S2). The overall variation was low, with a pooled standard deviation of just 0.615 (95% confidence interval: 0.504-0.790) and a categorical call concordance of 38/40 (95%). Both the discordant replicates were from a single biological sample that straddled the low to medium threshold. Two separate processed replicates did not pass QC and were not included in this analysis.

## DISCUSSION

Using the previously published OncoPrism biomarker capable of predicting ICI response in HNSCC patients (OncoPrism-HNSCC) we optimized and validated OncoPrism for NSCLC (OncoPrism-NSCLC).^21^ OncoPrism-NSCLC predicted PFS and OS in the OAK anti-PD-L1 ICI cohort and ORR, PFS, and OS in the 17 site PREDAPT anti-PD-1 ICI cohort (Figure 1C,D & Figure 3A,C,E). PD-L1 TPS failed to predict ORR, PFS, or OS in the PREDAPT anti-PD-1 cohort (Figure 3B,D,F). OncoPrism-NSCLC also demonstrated robust analytical performance with low variation across batches, operators, and kit lots (Figure S2).

Importantly, while OncoPrism-NSCLC predicted PFS and OS in the two ICI-treated patient cohorts, it did not predict PFS or OS in two non-ICI cohorts (*OAK non-ICI Cohort*, *TCGA non-ICI Cohort*) (Figure 5). This finding was consistent with previous observations with four non-ICI cohorts in the OncoPrism-HNSCC study, indicating that the biomarker underlying the OncoPrism-NSCLC test is predictive of ICI benefit rather than prognostic of outcome regardless of treatment, which would limit its clinical utility.^21^ Lack of predictive value in non-ICI patients also suggests that the predictive value of OncoPrism-NSCLC in chemo-immunotherapy treated patients may be derived from the ICI component of the regimen.

While the study met its primary endpoint of predicting ORR, there are several limitations and opportunities for future directions. Work is ongoing to recruit additional patients for subgroup analyses such as comparing performance in monotherapy ICI vs. chemo-immunotherapy treated patients. Because this cohort includes patients treated with ICI as a single agent and in combination with chemotherapy, there is a risk that treatment regimen biases the outcome, particularly ORR, across OncoPrism groups. Specifically, chemo-immunotherapy patients have a higher ORR than monotherapy patients (51% for chemo-immunotherapy therapy vs. 38% for monotherapy). However, there is no significant association between OncoPrism group and treatment regimen, indicating that the high ORR in OncoPrism high patients is not driven by treatment regimen (Figure 4A). Conversely, PD-L1 TPS≥50 is associated with treatment regimen, with an enrichment in monotherapy-treated patients (Figure 4A). Because PD-L1 TPS≥50 is used to select for monotherapy treatment, this association is unsurprising. PD-L1 TPS 1-49 and TPS≥50 have similar ORRs, which may be explained in part by the higher proportion of PD-L1 TPS 1-49 patients who received chemo-immunotherapy.

While significant, there is a relatively modest correlation between OncoPrism-NSCLC group and PD-L1 TPS category (Figure 4A-B), which may suggest potential complementary use of OncoPrism-NSCLC together with PD-L1 to improve predictive accuracy relative to either test alone. Similarly, future work will interrogate any relationship between OncoPrism-NSCLC and mutations in genes shown to be correlated with poor response to ICI (KEAP1 and STK11) and whether there is opportunity to further improve performance.^14^

Overall, OncoPrism-NSCLC is a laboratory developed test (LDT) that fulfills the current need for better predictive tests for NSCLC patients being considered for ICI. As reported here, OncoPrism-NSCLC is: (a) compatible with limited tissue from FFPE tumor biopsies; (b) robust against variation in the context of testing variables; (c) highly specific with good sensitivity, especially compared to PD-L1 TPS; and (d) predictive rather than prognostic, providing opportunity for greater clinical utility.

## METHODS

### Datasets

#### TCGA non-ICI Cohort (n=1,010) // Biomarker Distribution + Prognostic Evaluation

RNA-seq counts and clinical data for TCGA disease codes LUAD and LUSC were downloaded using TCGAbiolinks.^25^ We filtered for primary tumor samples and filtered out technical replicates such that only one sample per patient made it into the final dataset. This resulted in a final set of n=1,010.

#### OAK non-ICI Cohort (n=193) // Prognostic Evaluation

Access to the OAK trial (NCT02008227) clinical sequencing data and clinical outcome data was granted by Genentech.^23^ This cohort represented the arm of the trial treated with docetaxel 75 mg/m² (non-ICI) every 3 weeks until progressive disease or unacceptable toxicity. Two-thirds of the data was randomly selected for use, with one-third withheld for potential future analysis. This resulted in a final set of n=193.

#### OAK ICI anti-PD-L1 Cohort (n=195) // Predictive Evaluation

Access to the OAK trial (NCT02008227) clinical sequencing data and clinical outcome data was granted by Genentech.^23^ This cohort represented the arm of the trial treated with atezolizumab 1200 mg (ICI/anti-PD-L1) every 3 weeks until progressive disease or unacceptable toxicity. Two-thirds of the data was randomly selected for use, with one-third withheld for potential future analysis. This resulted in a final set of n=195.

#### PREDAPT ICI anti-PD-1 Cohort (n=89) // Predictive Evaluation

PREDAPT samples were collected and data generated as described in detail in other sections of these methods.

### Biomarker Distribution and Tertile Score Determination (OncoPrism-NSCLC)

The distribution of OncoPrism scores across six cancer types was determined using TCGA datasets for LUAD plus LUSC, KIRC, HNSC, COAD, the triple-negative breast cancer subset of BRCA, and BLCA (Figure 1A). Because the distribution of OncoPrism scores varied across cancer types, an NSCLC-specific distribution was created using a cohort of 1,010 TCGA (LUAD plus LUSC) patient samples. The resulting distribution was divided into tertiles to establish thresholds for all subsequent validations.

### PREDAPT Study Design and Participants

Patients were recruited from the following 17 academic and community study sites across the United States, with the aim of a representative sample of the affected population: Appalachian Regional Healthcare, Inc (Hazard, KY), Cancer Care Northwest (Spokane, WA), Cox Medical Centers (Springfield, MO), Gundersen Medical Foundation (La Crosse, WI), John B Amos Cancer Center (Columbus, GA), Lahey Hospital and Medical Center (Burlington, MA), MultiCare Institute for Research and Innovation (Tacoma, WA), Ochsner Lafayette General Medical Center (Lafayette, LA), Providence Regional Cancer System (Lacey, WA), Touro Infirmary LCMC Health (New Orleans, LA), Valley Cancer Associates (Harlingen, TX), William Beaumont Army Medical Center (Fort Bliss, TX), Brooke Army Medical Center (Fort Sam Houston, TX), Decatur Memorial Hospital (Decatur, IL), Holy Cross Hospital (Fort Lauderdale, FL), Northwest Oncology and Hematology (Hoffman Estates, IL), Sharp Clinical Oncology Research (San Diego, CA).

Patients were enrolled from 2021-2025 in a retrospective, observational study. No patient-level study data were reported to patients or physicians and the patients and public were not involved in the study design. Patients were enrolled following the inclusion and exclusion criteria outlined below. Eligible patients were adults with stage IV non–small cell lung cancer (NSCLC) at diagnosis who had received at least two doses of anti–PD-1 immunotherapy (pembrolizumab) as first-line single-agent treatment (monotherapy) or in combination with chemotherapy (chemo-immunotherapy therapy). Patients were required to have an Eastern Cooperative Oncology Group (ECOG) performance status of 0–2 prior to initiation of anti–PD-1 therapy as a single agent or 0-1 prior to initiation of anti-PD-1 therapy in combination with chemotherapy; if ECOG status was unavailable, documentation of self-care capacity and/or ambulatory activity for at least 50% of waking hours was accepted for monotherapy patients, or documentation of no requirement for assistance with daily living or activities for chemo-immunotherapy therapy patients. A pretreatment tumor biopsy was required, with at least 45µm FFPE tissue available for analysis. Evaluation of tumor response to therapy by imaging and/or clinical assessment must have been performed, typically 1-4 months after initiation of therapy.

Key exclusion criteria included a history of stage III or later non-NSCLC malignancies, recurrent non-NSCLC cancers, progression from early-stage (stage I–III) NSCLC, receipt of fewer than two cycles of anti–PD-1 therapy, or known targetable EGFR or ALK mutations. Patients who had previously received non-hormonal systemic therapy (e.g., platinum-based chemotherapy, targeted therapy, or immunotherapy) were excluded; prior hormonal therapy such as tamoxifen was permitted.

Tissue specimens analyzed in the study were collected from pre-treatment tumor samples originally processed as FFPE specimens using standard histologic protocols. De-identified, pre-treatment FFPE tumor biopsy specimens were provided to Cofactor Genomics for OncoPrism-NSCLC and PD-L1 IHC analysis. Following treatment, each patient’s tumor response to immunotherapy was evaluated using RECIST, PERCIST, or other clinical criteria as appropriate in standard of care to determine disease control. Patients with insufficient tissue for analysis or less than 10% neoplastic cellularity as determined by a study pathologist (EJD) were excluded from the study. Primary or metastatic tumor specimens were accepted, but metastatic tumors from brain, liver, or bone were not included. Length of follow-up ranged from 47 days to 68 months. The study protocol, “A Multicenter Cancer Biospecimen Collection Study” was registered on August 10, 2020 as “NCT04510129—PREDicting immunotherapy efficacy from Analysis of Pre-treatment Tumor biopsies (PREDAPT)” on clinicaltrials.gov. The study protocol was approved by institutional review boards at either the study (Advarra, Inc. [Columbia, MD] or WCG IRB [Puyallup, Washington]) or site level, as appropriate. All patients provided signed, informed consent to participate, or consent was waived for deceased patients according to study protocol. Independent data monitoring was conducted by the study clinical research organization Curebase, Inc. (San Francisco, CA).

### RNA Extraction

RNA was extracted using RNAstorm (Biotium, Fremont, CA) or the Zymo Quick-DNA/RNA FFPE Miniprep Kit (Irvine, CA) according to the manufacturer’s instructions for RNA isolation. RNA quantity was assessed by the High Sensitivity RNA Qubit assay (Thermo Fisher Scientific, Waltham, MA). A predefined yield of 20 ng FFPE RNA was used as the minimum QC threshold. Quality of the RNA was assessed using a bioanalyzer (Agilent Technologies, Santa Clara, CA), and a DV200 of at least 21% was required for all samples.

### Library Preparation and Sequencing

Libraries were prepared using the QuantSeq 3’ mRNA-Seq Library Prep Kit FWD for Illumina with the QuantSeq-Flex Targeted RNA-Seq Module (Lexogen, Inc., Greenland, NH) following the manufacturer’s instructions. Library RNA input was 20-40 ng. UMI Second Strand Synthesis Module for QuantSeq FWD (Lexogen, Inc., Greenland, NH) replaced Second Strand Synthesis Mix 1 in the workflow. All samples were processed with two OncoPrism positive controls and a No Template Control. The positive (high or medium scoring) controls were RNA extracted from RM-HNSCC FFPE samples.^23^ Final libraries were sequenced to a minimum depth of 5 million single-end 75 base pair reads on a NextSeq500 (Illumina, San Diego, CA), following the manufacturer’s protocols.

### Immunohistochemistry

PD-L1 staining and TPS was performed by NeoGenomics Laboratories (Fort Myers, FL) using the PD-L1 22C3 FDA for NSCLC stain. H&E staining was performed by NeoGenomics as part of the PD-L1 22C3 test or at Cofactor Genomics using xylene substitute Slide Brite (Newcomer Supply, Middleton, WI), as detailed by manufacturers. Neoplastic cellularity was estimated by a trained pathologist (EJD).

### Processing of RNA sequencing data

FASTQ files were preprocessed with trim_galore/cutadapt version 0.4.1 to remove adapter sequences, reads with PHRED quality scores less than 20, and reads shorter than 20 basepairs. The trimmed reads were aligned to the human genome GRCh38 with STAR version 2.5.2a using the two-pass method as previously described.^26^ Read counts were generated using htseq-count version 0.9.1 and annotation from Gencode version 22.^26^ The data was normalized as counts per million (CPM) and log2 transformed using unique reads aligning to protein coding regions. Samples were required to have a minimum of 30% exonic alignment and 800,000 unique deduplicated counts to be included in the study.

### Validation of PREDAPT Cohort Performance

Clinical validation of the OncoPrism-NSCLC test was performed using a separate cohort of 89 unique patient samples. Samples were processed in the Cofactor Genomics College of American Pathologists (CAP)-accredited, Clinical Laboratory Improvement Amendments (CLIA)-certified laboratory using strict quality controls. The primary validation metric was ORR in each OncoPrism Group. ORR was calculated by dividing the sum of patients with RECIST 1.1-defined categories of partial response and complete response as initial response by the total number of patients in each group. RECIST label was determined one to four months after initiation of ICI treatment when possible, but one patient was evaluated at six months due to treatment regimen and availability for follow-up imaging. Operators were blinded to the RECIST label when processing samples and generating OncoPrism scores. The RECIST labels for each patient were determined independently from the OncoPrism group and were used to determine the ORR for each group in the validation set. Patients with complete response or partial response were treated as the positive class (“responders”). Patients with stable disease or progressive disease were treated as the negative class (“non-responders”).

### Statistics

The primary endpoint of this study was overall response rate. A two-sided Cochran-Armitage test for trends was used to test the significance of the trend of increasing proportions for the ORRs of OncoPrism groups. Power analysis was performed using a range of potential AUCs of 0.68-0.72 and assuming a response rate of 50%. Under these assumptions, minimum cohort sizes of 60-100 were calculated for 80% power at p<0.05 for the primary endpoint (Cochran-Armitage Test for trend in proportions for ORR). For PREDAPT patients PFS was defined as the time from start of ICI treatment to progression or death. OS was defined as the time from start of ICI treatment to death by any cause. Patients were censored if they had not progressed at last follow up. PFS and OS figures and analysis were done using the “survminer” and “survival” packages, and significance was determined using log rank methods.^27,28^ 95% Confidence intervals for model performance metrics were calculated using a non-parametric bootstrap resampling method. Pairwise associations among categorical variables were tested using a chi-squared test of independence. In all cases a p-value of less than 0.05 was considered significant.

## DATA AVAILABILITY

All the relevant data supporting the findings of this study are provided within the main manuscript and its supplementary files. TCGA sequencing data is available from the TCGA consortium (https://www.cancer.gov/tcga). OAK sequencing data is obtained by request from Genentech through the European Genome-Phenome Archive (https://ega-archive.org/datasets/EGAD00001007703). PREDAPT sequencing data are derived from clinical samples. Due to IRB, hospital contracts, and HIPAA requirements, we are not authorized to share individualized patient genomic data. Cofactor Genomics is dedicated to collaboration and the advancement of science. Reasonable data requests outside the scope of this manuscript should be addressed to the corresponding author.

## ACKNOWLEDGMENTS

We would like to thank the patients who participated in this study. In addition, we thank all members of the PREDAPT study consortium. In particular, we thank the following people for their invaluable contributions: Sidney Aguirre, Rachel Allen, Uriah Sam Atkinson, Nowsheen Azeemuddin, Rajesh Bande, Rachel Bender, Darcie A. Cruz, Jo Ellyn Curran, Breanna N. Davis, Deirdre L. Dillon, Cheryl L. Dodd, Eileen M. Georgi, Richard Goforth, Leah A. Guilford-Elenes, Peter Jiang, Mukesh Kumar, Amber N. Larimer, Jenna R. Larry, Nagabhishek Moka, Elyssa Navarro, Uyen Nguyen, Suresh Nukala, Lacey Osborne, Denise Pichardo, Meley L. Pine, Christina Renshaw, Julie Roache, Alaina B. Sandoz, Julie Skarsvog, Galen A. Steinhoff, Dennis Spicer, Alierykel Williams, Erin L. Wilson, and Anna Young. The results shown here are in part based upon data generated by the TCGA Research Network: https://www.cancer.gov/tcga. Additionally, we would like to thank Genentech for providing access to their OAK clinical PD-L1 inhibitor clinical trial data (both the ICI-treated and chemotherapy-treated cohorts).

## AUTHOR CONTRIBUTIONS

Conception and design: KCF, JE, JH, RLW, MMP, KP, JAH, HLMcL, EJD, JIG. Recruitment of patients and acquisition of samples and data: MMP, JIG, OM, KW, AWP, KD, XS, WLA, JS, SJS, TDS, SDB, SAS, GA, BB, SEK, PG, JWIII.

Execution of the research: KCF, JE, JH, RLW, MMP, KP.

Analysis and interpretation of data: KCF, JE, JH, JIG.

Drafting of the manuscript: KCF, JE, JIG.

Guarantor: JIG.

---

**Figure S1.**
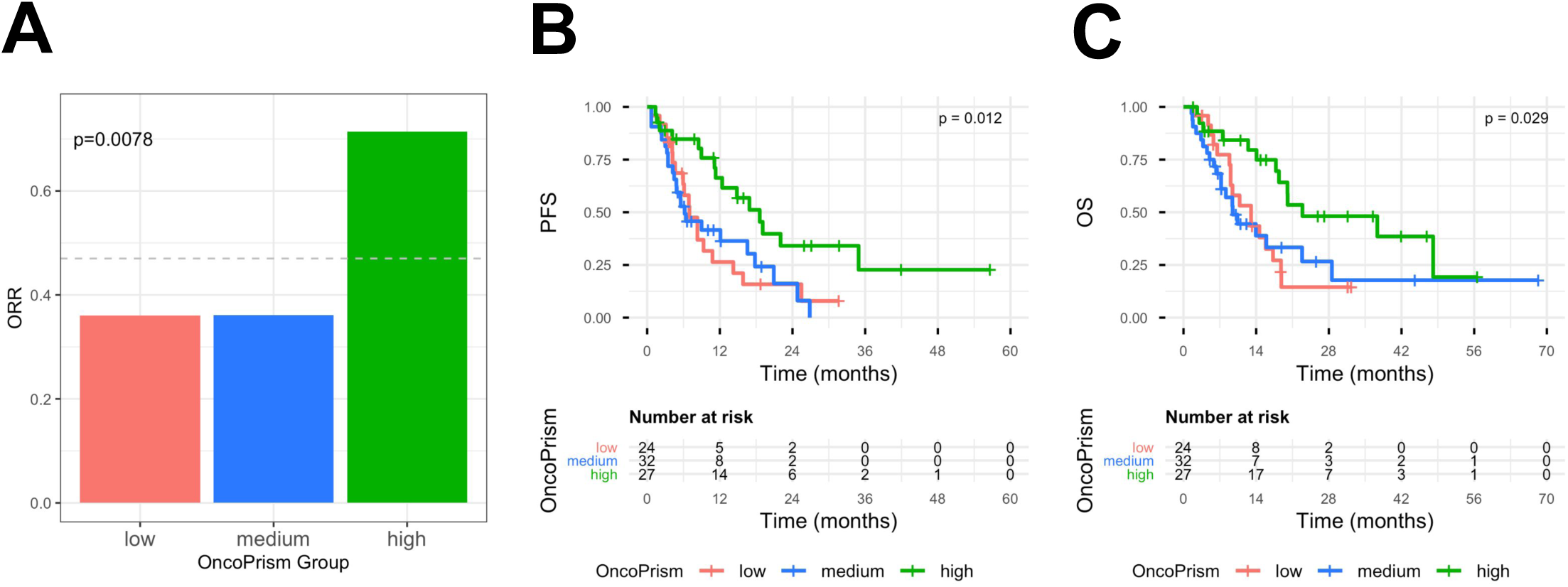
OncoPrism group is correlated with clinical benefit in the subset of PREDAPT patients where PD-L1 TPS data was available (n=83). **(a)** OncoPrism-NSCLC group was significantly correlated with overall response rate (ORR). The ORR for the entire n=83 cohort is shown as a dashed line. **(b)** OncoPrism-NSCLC significantly predicted progression-free survival (PFS). **(c)** OncoPrism-NSCLC significantly predicted overall survival (OS).

**Figure S2.**
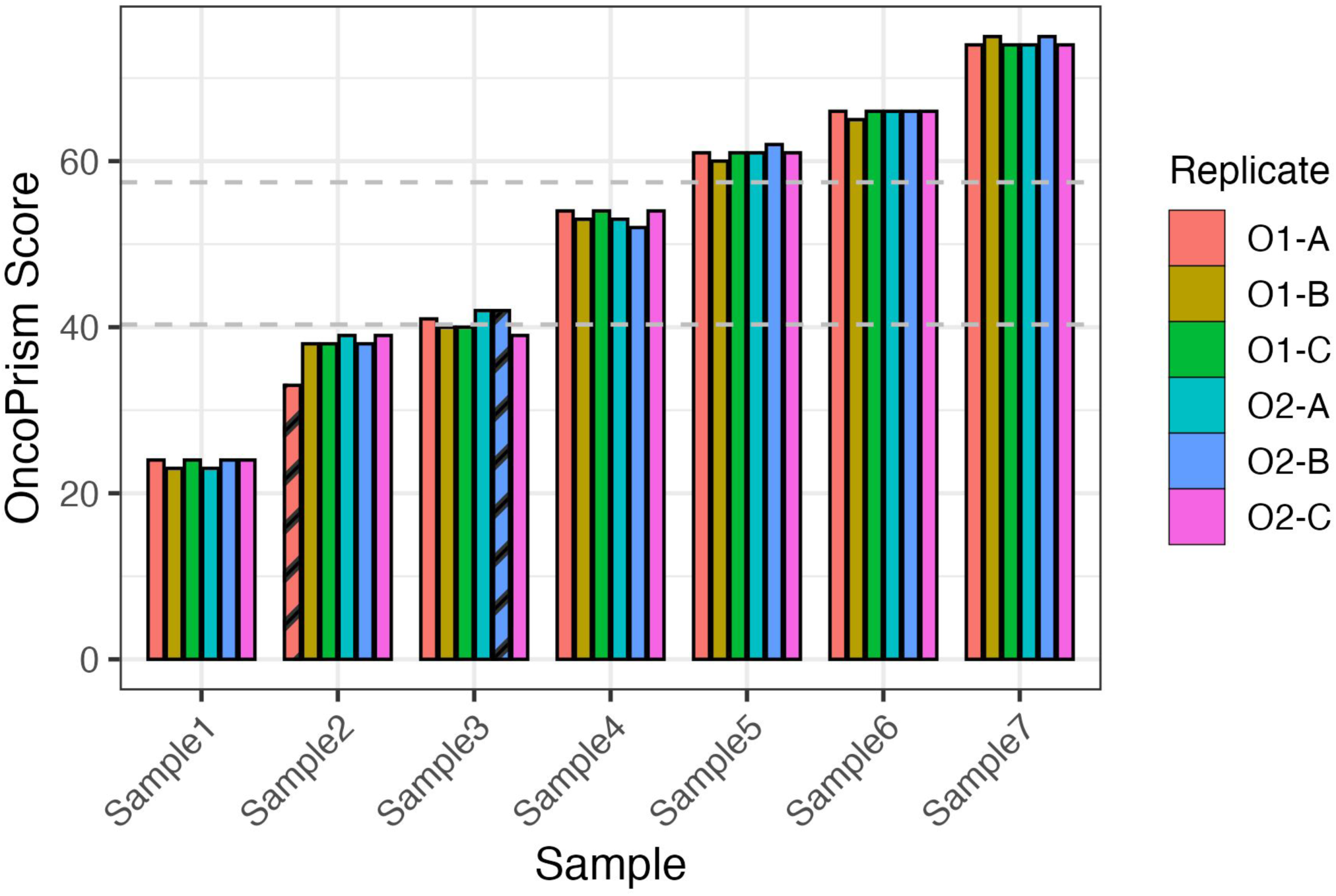
OncoPrism-NSCLC is analytically robust. Seven biological samples were processed in triplicate by two operators on two different days with two different reagent lots for each batch. Striped boxes represent replicates that did not pass QC. OncoPrism group thresholds are shown as dashed lines. “O1” is Operator 1. “O2” is Operator 2.

**Table S1.** Excluded Patient Samples.

|  |  |
| --- | --- |
| None (included in study) | 89 |
| Unacceptable Treatment Regimen | 5 |
| Other Concurrent Cancer | 2 |
| Unacceptable Specimen Site | 14 |
| Incomplete Data | 5 |
| Insufficient Material | 19 |
| Not NSCLC diagnosis | 1 |
| Tumor cellularity | 7 |
| Fail RNA QC | 27 |
| Fail Library QC | 0 |
| Fail Analytical QC | 7 |
| Withdrawn | 0 |
| **Total** | **176** |

**Table S2.** Performance metrics for OncoPrism-NSCLC and PD-L1 TPS.

|  | **Accuracy** | **PPV** | **NPV** | **Sensitivity** | **Specificity** |
| --- | --- | --- | --- | --- | --- |
| **OncoPrism**<br>(Low+Medium<br>vs High) | 66.3% | 71.4% | 63.9% | 47.6% | 83.0% |
| **PD-L1**<br>(TPS<50 vs<br>TPS≥50) | 54.2% | 57.1% | 52.7% | 38.1% | 70.7% |

\*PPV=Positive Predictive Value; NPV=Negative Predictive Power

**Table S3.** OncoPrism and PD-L1 predicted and actual outcomes.

|  | PD-L1 TP | PD-L1 FN | PD-L1 TN | PD-L1 FP |
| --- | --- | --- | --- | --- |
| OncoPrism TP | 11 | 9 | - | - |
| OncoPrism FN | 5 | 17 | - | - |
| OncoPrism TN | - | - | 26 | 8 |
| OncoPrism FP | - | - | 3 | 4 |

\*TP=True Positive; FN=False Negative; TN=True Negative; FP=False Positive

